# Titers and capacity of neutralizing antibodies against SARS-CoV-2 variants after heterologous booster vaccination in health care workers primed with two doses of ChAdOx1 nCov-19: a single-blinded, randomized clinical trial

**DOI:** 10.1101/2022.06.14.22276236

**Authors:** Chih-Hsien Chuang, Chung-Guei Huang, Ching-Tai Huang, Yi-Ching Chen, Yu-An Kung, Chih-Jung Chen, Tzu-Chun Chuang, Ching-Chi Liu, Po-Wei Huang, Shu-Li Yang, Po-Wen Gu, Shin-Ru Shih, Cheng-Hsun Chiu

## Abstract

**Background:** Booster vaccination is important because of waning immunity and variant immune evasion. We conducted a single-blinded, randomized trial to evaluate the safety, reactogenicity, and immunogenicity of heterologous booster vaccination in health care workers (HCW) who had received two doses of ChAdOx1 nCov-19.

**Methods and findings:** HCW at least 90 days after the second dose were enrolled to receive one of the four vaccines: BNT162b2, half-dose mRNA-1273, mRNA-1273, and MVC-COV1901. The primary outcomes were humoral and cellular immunogenicity and the secondary outcomes safety and reactogenicity 28 days post-booster. 340 HCW were enrolled: 83 received BNT162b2 (2 excluded), 85 half-dose mRNA-1273, 85 mRNA-1273, and 85 MVC-COV1901. mRNA vaccines had more reactogenicity than protein vaccine.

Anti-spike IgG increased by a fold of 8.4 for MCV-COV1901, 32.2 for BNT162b2, 47.6 for half-dose mRNA-1273 and 63.2 for mRNA1273. The live virus microneutralization assay (LVMNA) against the wild type, alpha and delta variants were consistent with anti-spike IgG for all booster vaccines. The LVMNA in the four groups against omicron variant were 6.4 to 13.5 times lower than those against the wild type. Serum neutralizing antibody against omicron variant was undetectable in 60% of the participants who received MCV-COV1901 as a booster by LVMNA. By using pseudovirus neutralizing assay, we found that neutralization activity in the four groups against omicron variant were 4.6 to 5.2 times lower than that against the D614G. All booster vaccines induced comparable T cell response.

**Conclusions:** Third dose booster not only increases neutralizing antibody titer but also enhances antibody capacity against SARS-CoV-2 variants. mRNA vaccines are preferred booster vaccines for those after primary series of ChAdOx1 nCov-19.

**Trial registration:** ClinicalTrials.gov NCT05132855

## Introduction

The pandemic of Coronavirus Disease 2019 (COVID-19) has been shaped by the successive emergence of different SARS-CoV-2 variants with increased transmissibility and/or immune escape, compared to the ancestral strain, since 2020 [1,2]. Delta (B.1.617.2) emerged in India in February 2021 and became dominant over the following months [2,3]. Omicron (B.1.1.529) that carries a large number of mutations in the spike protein gene was first reported from Gauteng province, South Africa in November 2021 [4]. Because the mutations are the major target of neutralization antibodies, omicron is able to avoid neutralization by serum from vaccinated or recovered individuals as well as by a large range of human monoclonal antibodies in use [1,5□7].

Taiwan experienced its first large wave of COVID-19 caused by alpha variant (B.1.1.7) from May to August 2021. The COVID-19 vaccination program in Taiwan started in March 2021. However, the first large batch of vaccines that arrived in Taiwan was ChAdOx1 nCov-19 (AstraZeneca). Majority of the health care workers (HCW) in Taiwan received two doses of ChAdOx1 nCov-19; first and second doses were given 8 weeks apart.

Previous studies have shown that effectiveness of two doses of ChAdOx1 nCov-19 vaccine against symptomatic disease among persons with alpha or delta was less than that of two doses of BNT162b2 vaccine (Pfizer BioNTech) [3]. Moreover, given the occurrence of rare, but severe adverse events after vaccination with vector-based vaccines such as ChAdOx1 nCov-19, heterologous prime-boost regimens have been recommended in many countries [8,9]. On the other hand, significant waning of humoral responses within 6 months after receipt of the second dose of BNT162b2 vaccine or inactivated CoronaVac vaccine (Sinovac Life Sciences) has been observed in recent studies [10,11]. Vaccine effectiveness in preventing COVID-19 might decline because of such waning immunity and furthermore, variant immune evasion. It is therefore critical to give a third dose, also known as a booster, to protect the vulnerable persons, and mitigate health care and economic impacts.

“Mixing and matching” COVID-19 vaccines may enhance the flexibility of vaccination and induce broader immune responses [12]. Immunological and safety assessments of the mRNA and Ad26.COV2.S boosters in persons who have received different priming regimens were reported before [13]. Protein vaccines are considered having higher safety than mRNA vaccines. MVC-COV1901 is a CpG1018 and aluminum hydroxide-adjuvanted SARS-COV-2 pre-fusion-stabilized spike protein S-2P vaccine developed by Medigen Vaccine Biologics Corporation, Taiwan. After a large phase II trial demonstrating good safety profile and promising immunogenicity, it was authorized for emergency use by Taiwan Food and Drug Administration (FDA) in July 2021. MVC-COV1901 is one of the two vaccines for inclusion in the WHO Solidarity Trial. To support decision making for persons in Taiwan and other countries who have received primary immunization with two doses of ChAdOx1 nCov-19, we performed the single-blinded, randomized study for a head-to-head comparison of safety and immunogenicity of different heterologous boosters administered to HCW.

## Methods

### Study design

The trial is to investigate the safety, reactogenicity and immunogenicity of heterologous boost dose of either BNT162b2, half-dose mRNA-1273 (Moderna), mRNA-1273 or MCV-COV1901 COVID-19 vaccine among HCW in a single institute at Chang Gung Memorial Hospital, Taoyuan, Taiwan. All participants gave written informed consent before entering this trial. The study protocol was approved by Chang Gung Medical Foundation Institutional Review Board (202101767A3).

### Participants

Participants were between 20 to 65 years of age and had received 2 doses of ChAdOx1 nCoV-19 vaccine for more than 90 days before enrollment. The main exclusion criteria were history of laboratory confirmed COVID-19 and anaphylaxis or severe allergic reaction to any components of study vaccines. The detail inclusion and exclusion criteria are in the S1 Text.

### Randomization and masking

Participants were randomly assigned, in a 1:1:1:1 ratio, to receive a single dose of BNT162b2 (30μg), half-dose mRNA-1273 (50μg), mRNA-1273 (100μg) or MVC-COV1901 (15μg). Computer generated randomization list with block sizes of four was used for randomization. Participants were blinded to boost vaccine until 180 days postvaccination. Laboratory staff processing immunological samples were blinded to vaccine allocation. Clinical nurses and research staff accessing adverse events were unblinded.

### Procedures

After an online screening procedure, participants were invited to join the study. Baseline demographic data were collected via electronic questionnaire. Blood samples were taken for baseline hematological and biochemical testing before vaccination at first visit. Blood samples were collected for immunogenicity analysis and checking for any previous infection before vaccination and 28 days after vaccination. We accessed adverse events by use of a modified US FDA toxicity grading scale [14]. Participants were asked to record electronic questionnaire on solicited local and systemic adverse events daily for 7 days and unsolicited adverse events weekly for 28 days post-booster. Serious adverse events and adverse events of special interest were reported by telephone or mobile message app for 180 days after booster vaccination.

### Immunogenicity

All blood samples were measured for quantitative anti-spike IgG, SARS-CoV-2 nucleocapsid IgG, surrogate neutralizing antibody by an ELISA kit (Formosa Biomedical Technology Corporation), and T-cell ELISpot kit. Serum anti-spike IgG concentrations were evaluated by use of Abbott AdviseDx SARS-CoV-2 IgG II assay [15]. SARS-CoV-2 nucleocapsid IgG were measured for confirmation of previous infection with the use of Roche Elecsys Anti-SARS-COV-2 [16]. The MeDiPro SARS-CoV-2 Antibody ELISA kit, based on the binding affinity of S1 and RBD domains to antibodies, was designed to indirectly quantify SARS-CoV-2 neutralizing antibodies in the serum [17]. Values <12 IU/mL were considered negative. The kit was approved by the Taiwan FDA (No. 1106803303). SARS-CoV-2 spike protein-specific T cell response was evaluated by ex vivo stimulating peripheral blood mononuclear cells with use of the Human IFN-γ ELISpot Kit (EL285, R&D) [18]. All these kits were used following manufacturers’ instructions.

A random subset of 120 participants (30 in each group) was tested for live virus neutralization with wild-type, alpha, delta and omicron variants of SARS-COV-2, as well as pseudovirus neutralization with the pseudoviruses either expressing SARS-CoV-2 D614G or omicron spike protein. The live virus neutralizing antibody test followed the standard protocol of a live virus microneutralization assay (LVMNA) [19]. The lower limit of LVMNA was 34.45 IU/ml. SARS-CoV-2 pseudovirus expressing D614G or omicron spike protein were prepared and titrated by the National RNAi Core Facility, Academia Sinica, Taiwan. SARS-CoV-2 pseudovirus neutralization assay (PNA) was performed as previously described [20]. The lower limit of PNA was 8 ID_50_. Values below the limit were substituted with half of the cutoff value. The detail methods for the analysis of immunogenicity are available in the S2 Text Methods.

### Outcomes

The primary outcomes were immunogenicity assessed 28 days after booster vaccination, including serum SARS-CoV-2 anti-spike IgG concentration, the 50% neutralizing antibody titers (NT_50_) against wild-type, alpha, delta and omicron variants, and IFN-γ secreting T cells specific to whole spike protein of the wild type. Secondary outcomes were safety and reactogenicity including occurrence of solicited and unsolicited adverse events, adverse events of special interest and serious adverse events.

### Statistical analysis

The sample size was calculated on basis of previous report with a minimum clinically important difference of 1.75-times difference between geometric mean titer, assuming a SD of 0.4 on log_10_ scale [12]. Since six comparisons were performed between 4 groups, a significance level of 0.05/6=0.0083 was adjusted by Bonferroni correction. We need to recruit 64 participants in each group to achieve 90% power at a two-side 0.008 % significance level with 20% dropout rate. We enrolled 85 participants in each group. Categorical data are expressed as number (percentage). Continuous variables are presented as median (interquartile range). Immunogenicity endpoints were reported as point estimates with 95% confidence intervals. The fold change was calculated as the antilogarithm of the mean difference between the log10 transformed titer of post-boost and that of pre-boost. Analysis of differences in immunogenicity between booster vaccine groups were performed by use of analysis of variance with Scheffe post Hoc test. Correlations between different immunological tests were assessed by using Pearson correlation coefficient. Adverse events between groups were compared by Fisher’s exact test. *P* < 0.008 was considered statistically significant. All analysis was performed with SPSS statistical software version 21.

## Results

From Nov 29 to Dec 14, 2021, 852 participants were contacted, among whom 340 participants were enrolled and randomly assigned to 4 groups (Fig 1). No participants dropped out of the study. Two participants had detectable anti-nucleocapsid protein IgG antibody at baseline and during follow-up and were excluded for analysis. One participant had pregnancy after booster vaccine and was still under analysis. Among the 338 participants, the median age was 36 years old (range: 22-64) and 228 (67.5%) participants were female. The baseline demographic characteristics and laboratory tests were balanced across the 4 vaccine groups (Table 1). There were no differences in immunological studies including anti-spike IgG levels, surrogate neutralizing antibody levels by ELISA and S-specific T cell response at baseline across the 4 vaccine groups (Table 2).

**Fig 1.**
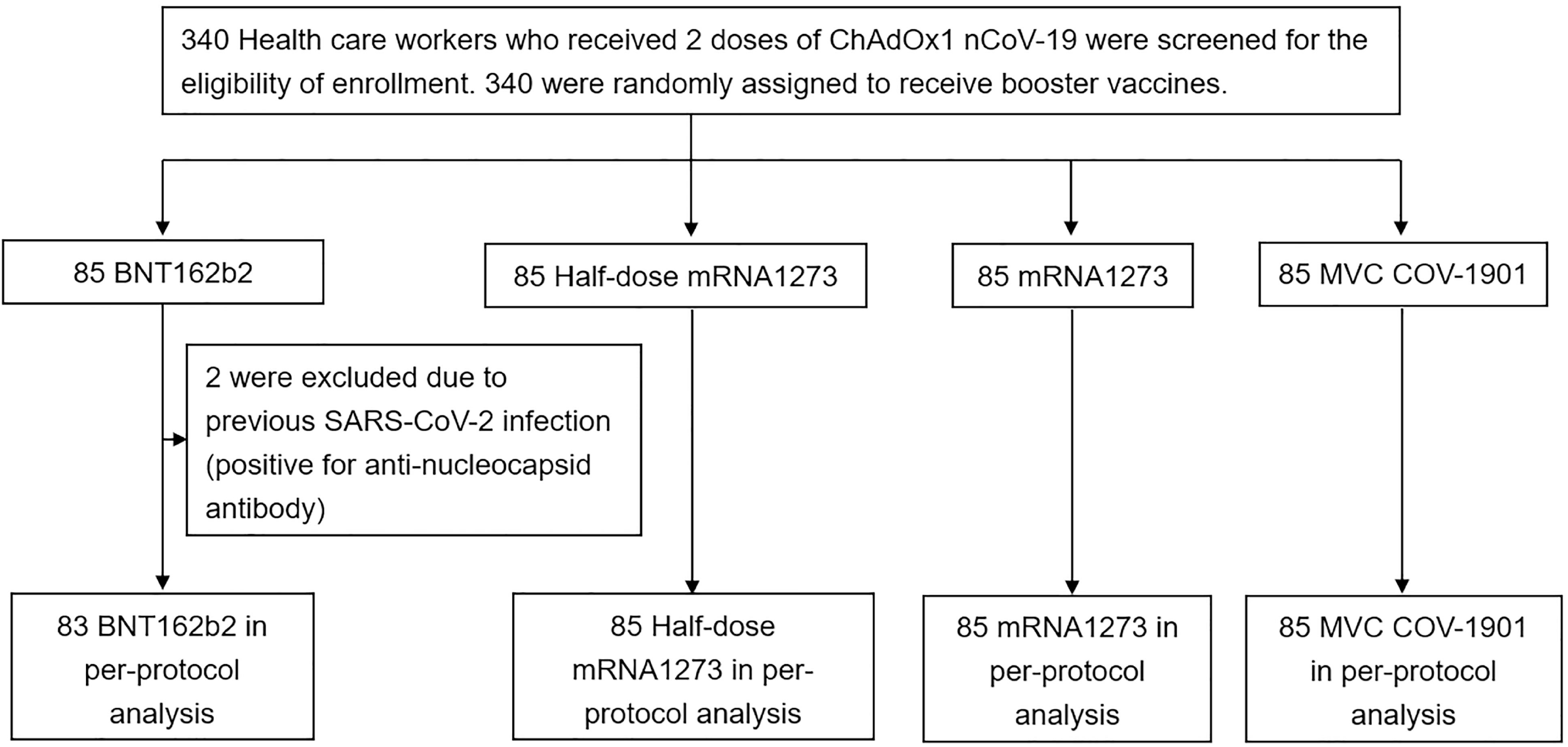
Study population and analysis. 340 participants were enrolled and randomly assigned to 4 groups. Two participants had detectable anti-nucleocapsid protein IgG antibody at baseline and during follow-up and were excluded for analysis.

**Table 1.** Baseline characteristics of the participants.

| **Characteristic** | **BNT162b2** | **Half-dose**<br>**mRNA-1273** | **mRNA-1273** | **MVC-COV1901** |
| --- | --- | --- | --- | --- |
| No. of participants | 83 | 85 | 85 | 85 |
| Sex – no. (%) |  |  |  |  |
| Female | 53 (64) | 58 (68) | 60 (71) | 57 (67) |
| Male | 30 (36) | 27 (32) | 25 (29) | 28 (33) |
| Age - years |  |  |  |  |
| Median (IQR) | 35.0 (30.0-44.0) | 35.0 (30.0-45.5) | 37.0 (30.5-44.0) | 39.0 (32.5-44.5) |
| Intervals between first and<br>second doses – days |  |  |  |  |
| Median (IQR) | 63 (59-71) | 68 (60-71) | 63 (61-71) | 68 (61-71) |
| Intervals between second doses<br>and booster – days |  |  |  |  |
| Median (IQR) | 138 (126-177) | 140 (128-182) | 138 (127-149) | 138 (131-177) |
| Body-mass index* | 22.6 (20.3-24.8) | 23.5 (21.9-26.3) | 23.1 (21.5-25.8) | 22.2 (20.1-25.5) |
| Comorbidities |  |  |  |  |
| Cardiovascular disease –no. (%) | 6 (7) | 2 (2) | 10 (12) | 6 (7) |
| Diabetes mellitus –no. (%) | 0 (0) | 1 (1) | 4 (5) | 2 (2) |
| Liver disease –no. (%) | 2 (2) | 3 (4) | 4 (5) | 2 (2) |
| Kidney disease –no. (%) | 0 (0) | 1 (1) | 0 (0) | 1 (1) |
| White blood cell count – per mm<sup>3</sup> |  |  |  |  |
| Median (IQR) | 5900 (5300-7200) | 6500 (5500-7850) | 6500 (5500-7750) | 6000 (5300-7700) |
| Hemoglobin – per g/dl |  |  |  |  |
| Median (IQR) | 13.8 (13.0-15.1) | 13.7 (13.0-14.7) | 13.5 (12.7-14.7) | 13.7 (12.8-14.6) |
| Platelet count – 10<sup>3</sup> per mm<sup>3</sup> |  |  |  |  |
| Median (IQR) | 272 (232-311) | 282 (240-334) | 278 (247-310) | 263 (233-311) |
| AST – U/liter |  |  |  |  |
| Median (IQR) | 18 (14-23) | 17 (13-20) | 17 (14-21) | 17 (14-21) |
| ALT – U/liter |  |  |  |  |
| Median (IQR) | 15 (10-24) | 15 (11-22) | 16 (11-23) | 16 (10-21) |

IQR= interquartile range. AST=aspartate aminotransferase. ALT=alanine aminotransferase.
\* Body-mass index is the weight in kilogram divided by the square of the height in meters.

**Table 2.**
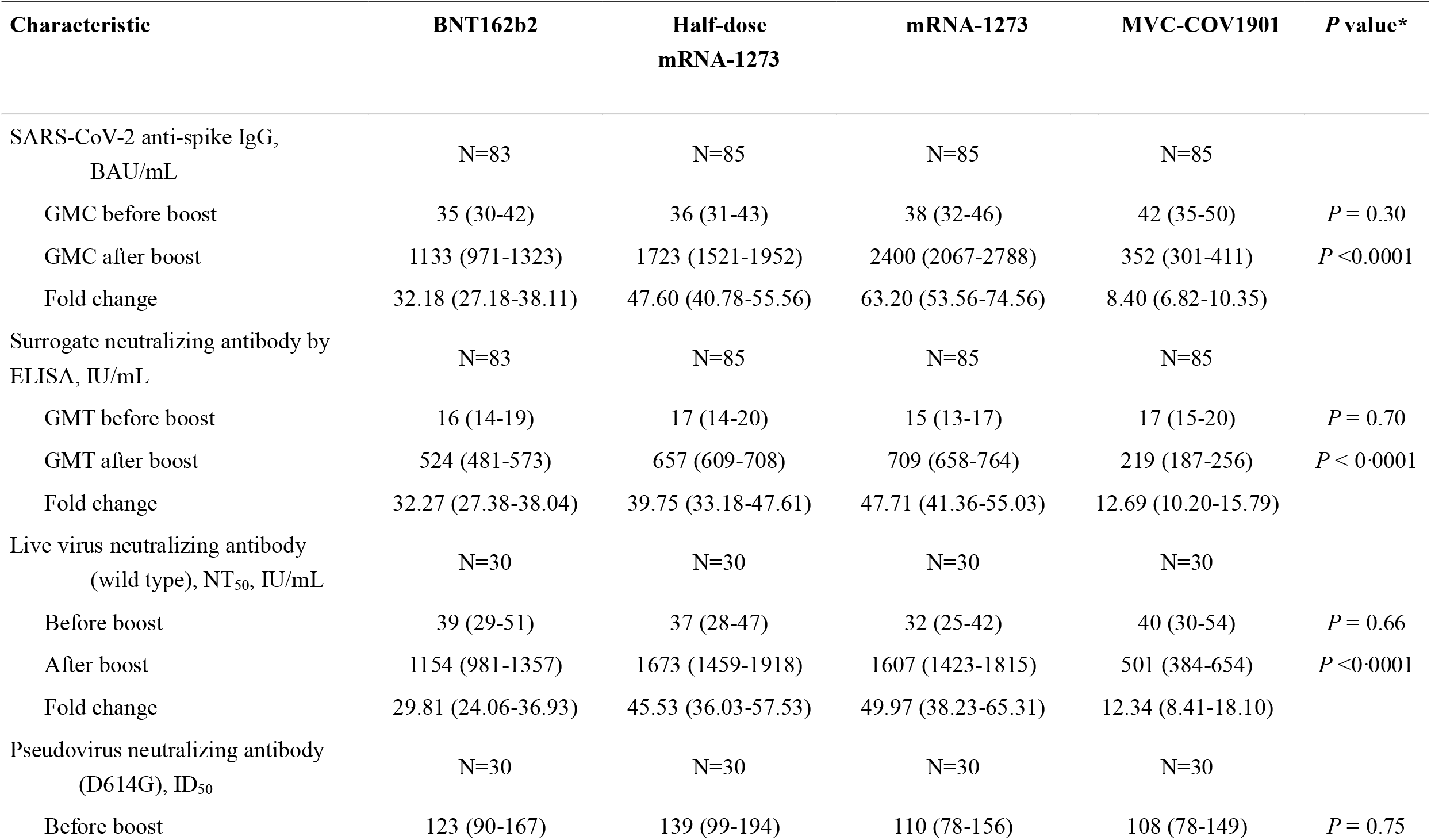
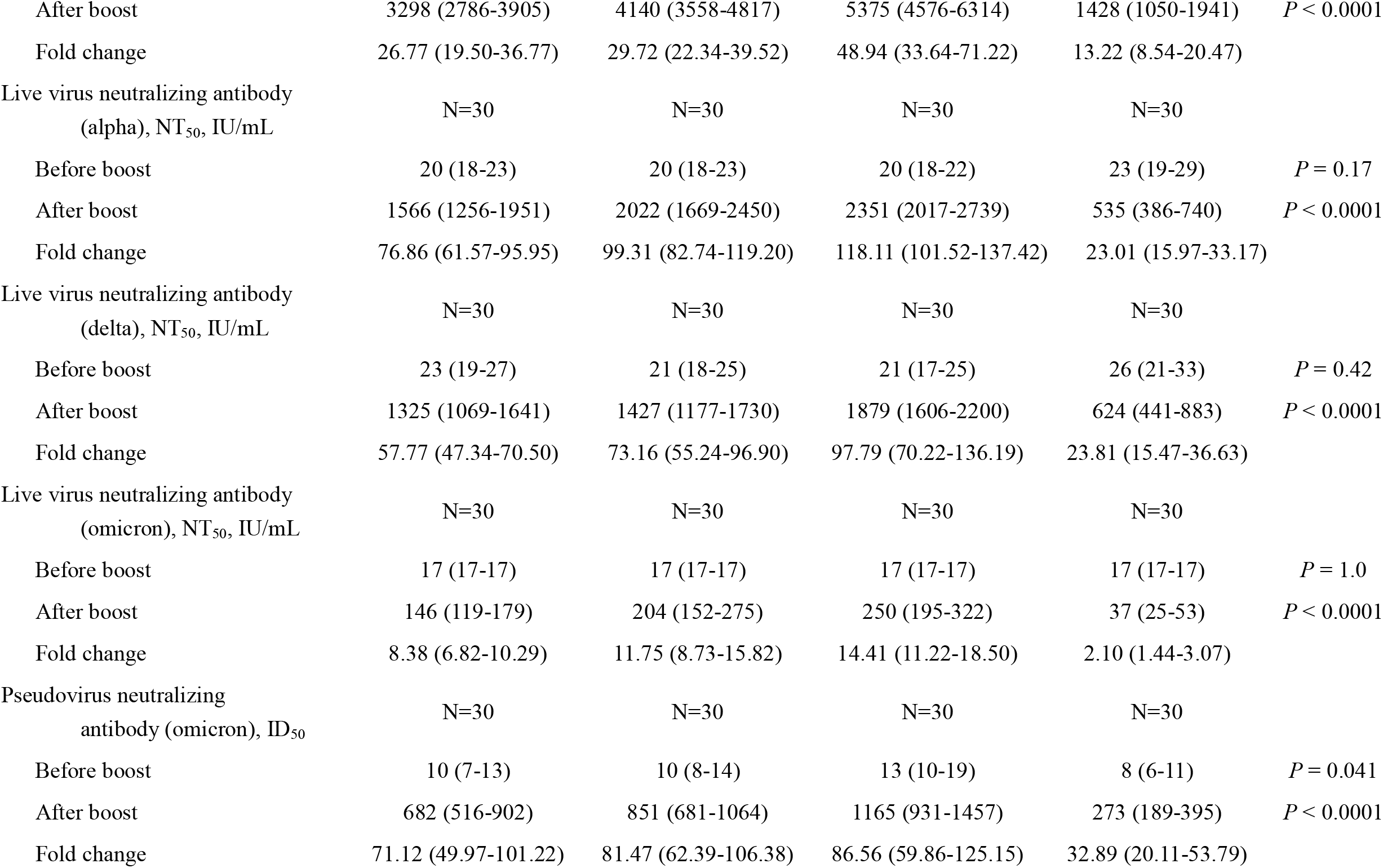
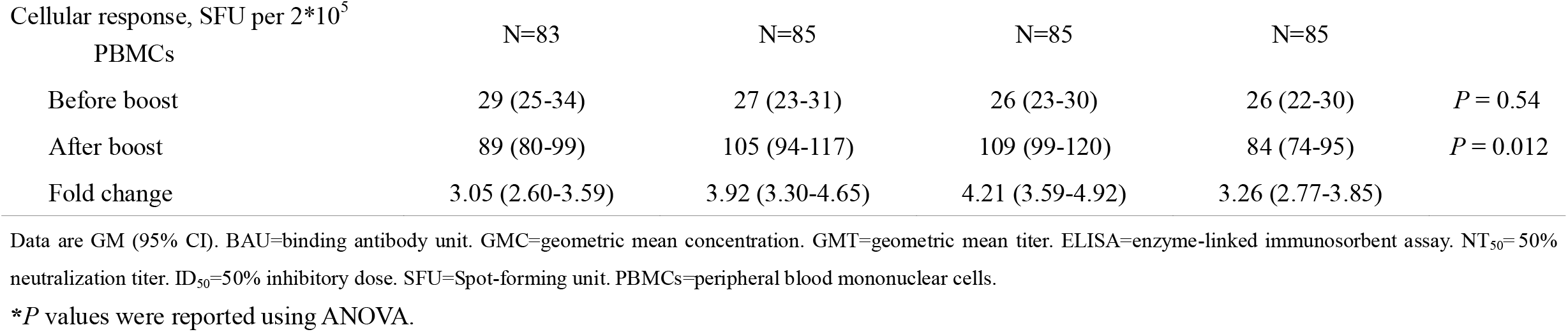
Binding antibody, neutralizing antibody, and T cell response.

No potential life threatening solicited or unsolicited adverse events were reported after any booster vaccine in the study. The most common local solicited adverse event was injection site pain (92%), followed by swelling (81%) and redness (13%). Fatigue (73%) was the most common systemic adverse event, followed by myalgia (67%), headache/dizziness (50%), nausea (21%), fever (15%), and diarrhea (15%) (see S1 Data Table A). The proportion of severe adverse events were injection site pain, 0 to 2.4%; fever, 0 to 5.9%; fatigue, 0 to 4.7%; myalgia, 0 to 1.2% and headache, 0 to 1.2% (Fig 2; S1 Data Table A). Local adverse events and some systemic adverse events such as fatigue, myalgia and headache could last for 1 week after booster vaccination (S2 Data Fig A). Fever subsided within 48 hours in most of the participants. Protein vaccine (MCV-COV1901) showed less local and systemic adverse events than mRNA vaccines. Full-dose mRNA1273 had more local and systemic adverse events than half-dose mRNA1273. Half-dose mRNA1273 had more local and systemic adverse events than BNT162b2 (Fig 2).

**Fig 2.**
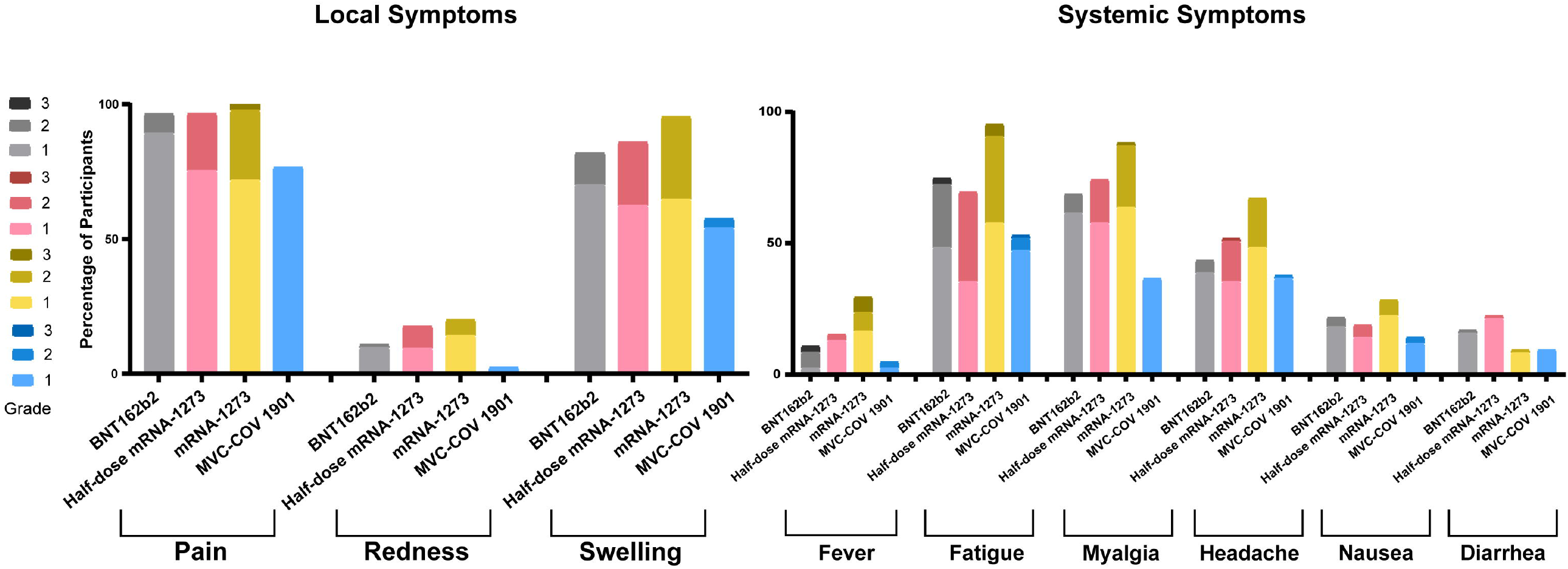
Severity of systemic and local reactions after booster vaccination. The percentage of participants with local symptoms (pain, redness, or swelling at the injection site) and the percentage of participants with systemic symptoms (fever, fatigue, myalgia, headache, nausea, or diarrhea) after booster vaccination are demonstrated. These reactions were monitored in the 7 days after the administration of the booster. The toxicity grading scale represents the highest grade of severity during the seven days. Grade 1 means mild reaction, grade 2 moderate reaction and grade 3 severe reaction. Grey: BNT162b2; red: half-dose mRNA-1273; yellow: mRNA-1273; blue: MVC-COV1901.

Compared with pre-boost, all study vaccines elicited significantly higher anti-spike IgG at 28 days post-boost (*P* < 0.0001). The fold rise ranged from 8.4 in MCV-COV1901 group to 63.2 in mRNA1273 group (Table 2). Significant differences were found in anti-spike IgG between study vaccines (*P* < 0.0001) (Fig 3). There were 262 (78%) participants who showed neutralizing antibodies below the detection limit by using surrogate neutralizing antibody ELISA kit before receiving booster vaccines. After booster vaccination, the fold rise ranged from 12.7 in MCV-COV1901 group to 47.7 in mRNA1273 group.

**Fig 3.**
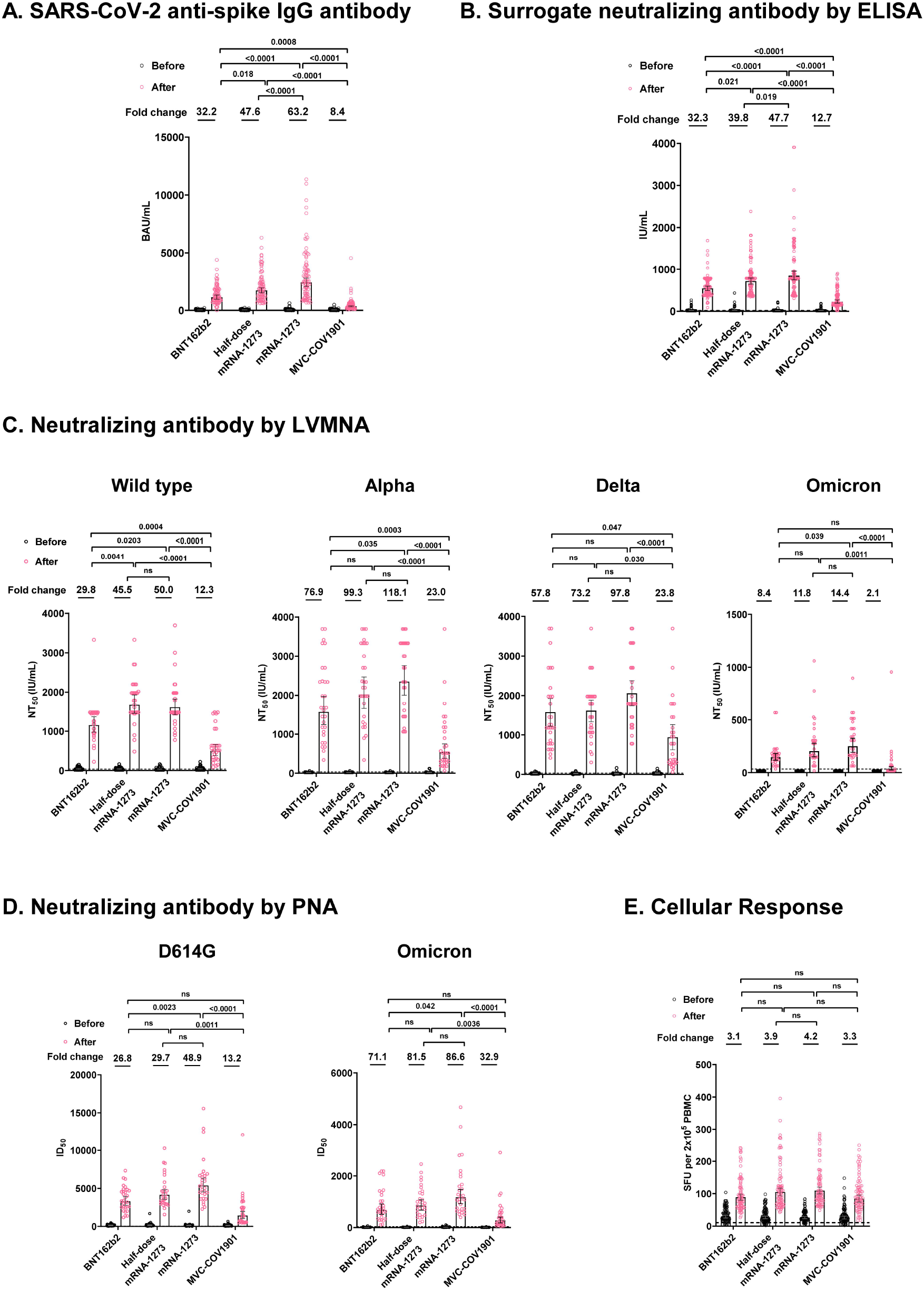
SARS-CoV-2 spike protein–specific immune responses before and after booster vaccination. Panel A shows levels of SARS-CoV-2 spike protein-specific IgG antibodies at baseline (before booster vaccination) and after booster vaccination in the four groups. Panel B shows levels of surrogate neutralizing antibodies by ELISA at baseline and after booster vaccination in the four groups. Panel C shows the levels of neutralizing antibodies at baseline and after booster vaccination, as assessed with a live virus microneutralization assay (LVMNA) in the four groups. Panel D shows the levels of neutralizing antibodies at baseline and after booster vaccination, as assessed with a SARS-CoV-2 pseudovirus neutralization assay (PNA) in the four groups. Panel E shows SARS-CoV-2 spike protein-specific T-cell response at baseline and after booster vaccination in the four groups, as measured by interferon-γ levels produced peripheral blood mononuclear cells after ex vivo stimulation. The *P* values on the top of figure were the comparison of the immune responses between groups.

The live virus neutralizing antibody against the wild type, alpha, delta and omicron variants pre-booster were undetectable in 48 (40%), 96 (80%), 87(73%) and 120 (100%) participants, respectively. The neutralizing activity increased significantly post boost (*P* < 0.0001), the fold-rise ranged from 12.3 in MCV-COV1901 to 50.0 in mRNA1273 against wild-type, 23.0 in MCV-COV1901 to 118.1 in mRNA1273 against alpha variant, 23.8 in MCV-COV1901 to 97.8 in mRNA1273 against delta variant and 2.1 in MCV-COV1901 to 14.4 in mRNA1273 against omicron variant (Fig 3). After each booster vaccine, the neutralization titers against alpha and delta variants were compatible with those of the wild type. The neutralization titers against omicron variant were 6.4 to 13.5 times lower than those against the wild type. All except one participant who received mRNA vaccines as a booster had detectable neutralizing antibody against omicron variant. Serum neutralizing antibody against omicron variant lower than the detection limit was observed in 60% of the participants who received MCV-COV1901 booster. The fold rise of pseudovirus neutralization against D614G ranged from 13.2 in MCV-COV1901 to 48.9 in mRNA1273. The pseudovirus neutralization against omicron variant were 4.6 to 5.2 times lower than that against D614G. In general, mRNA vaccines elicited significantly higher neutralizing antibody than protein vaccine. There was no significant difference in neutralizing antibodies between BNT162b2, half-dose mRNA1273 and mRNA1273. (Fig 3).

Level of interferon-γ was under the detection limit in 10 (3%) participants before boost. All vaccines induced significant T-cell response by ELISpot (Table 2). All booster vaccines elicited at least 3-fold rise of interferon-γ. There was no significant difference in T cell response between study vaccines.

Moderate correlations were found between anti-spike IgG and LVMNA against the wild type and variants of concern (VOCs) post boost (Pearson correlation coefficient of 0.53 to 0.64) (Fig 4). The correlations between different immunogenicity tests are provided in S2 Data Figs B□D.

**Fig 4.**
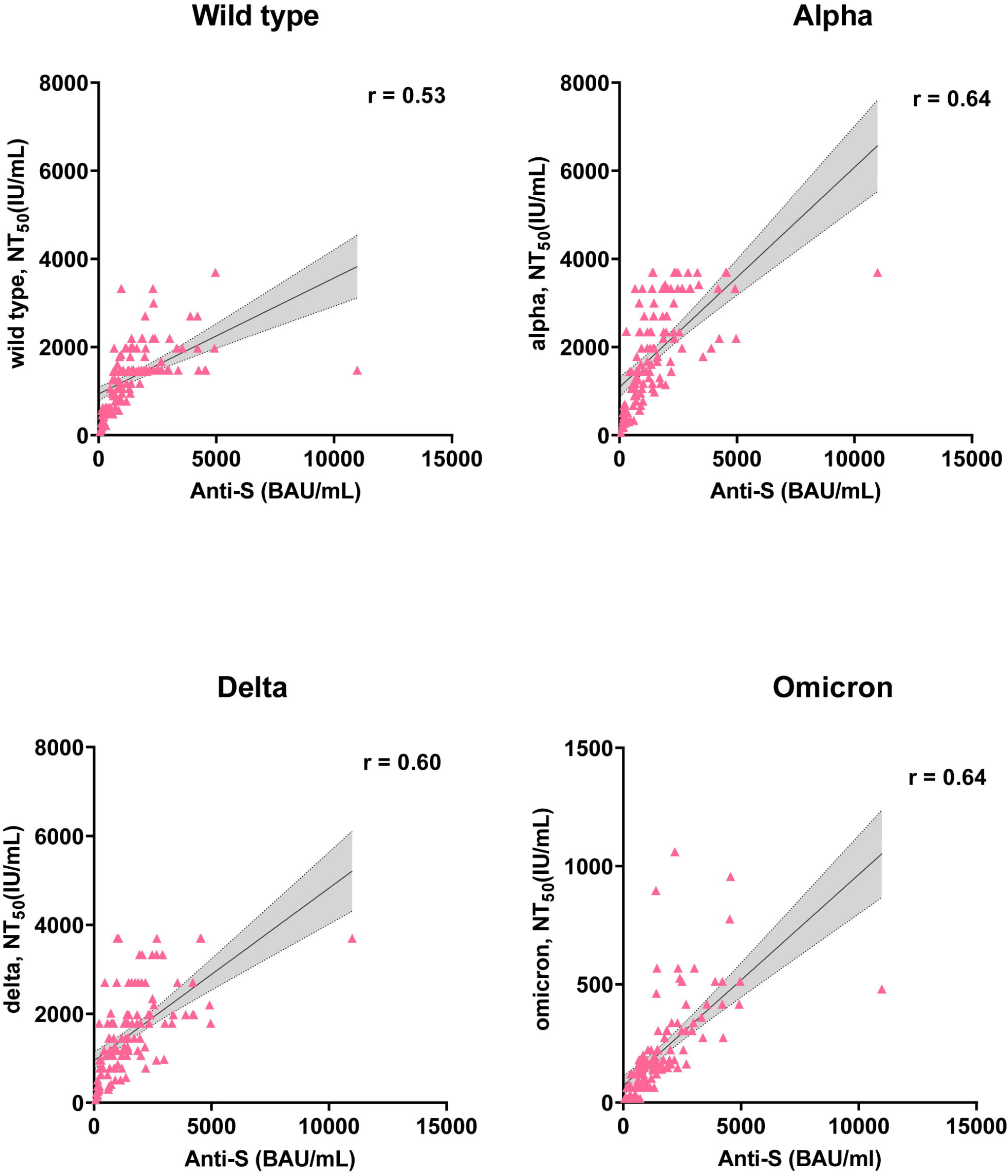
Correlation between anti-spike IgG and neutralizing antibodies after booster vaccination. The correlations between anti-spike IgG and neutralizing antibodies against the wild type by LVMNA (Pearson correlation coefficient, 0.53; *P* < 0.0001) (n = 120), neutralizing antibodies against the alpha variant by LVMNA (Pearson correlation coefficient, 0.64; *P* < 0.0001) (n = 120), neutralizing antibodies against the delta variant by LVMNA (Pearson correlation coefficient, 0.60; *P* < 0.0001) (n = 120), and neutralizing antibodies against the omicron by LVMNA (Pearson correlation coefficient, 0.64; *P* < 0.0001) (n = 120) are demonstrated. The gray shaded areas indicate the 95% CI of the best-fit line. Each dot in the figure represents an individual participant.

## Discussion

Data on protein vaccine as a booster and head-to-head comparison between different doses of mRNA vaccines remain scarce [21]. Our study addressed the immunogenicity, safety, and reactogenicity of currently recommended booster vaccines during the omicron pandemic. Furthermore, the prevalence of SARS-CoV-2 infection before the study in community in Taiwan is very low. Not affected by community-circulating virus, it is likely the immunogenicity shown in this study truly represents the vaccination effect.

The current study revealed that mRNA vaccines elicited more adverse events than protein vaccine and the reactogenicity was compatible to previous reports [12]. Compared with COV-BOOST [12], using BNT162b2 or mRNA1273 as a booster caused slightly more local and systemic adverse events in persons primed with 2 doses of ChAdOx1 nCov-19 in this study. MVC-COV1901 booster showed similar adverse event patterns to NVX-CoV2373 booster, except for local swelling and myalgia.

The antibody response of BNT162b2 and mRNA1273 boosting was generally consistent with previous reports from UK and USA [12,13,22]. The serum level of anti-spike IgG and live virus neutralization activity induced by MVC-COV1901 was approximately 31% to 47% of the response by BNT162b2. The antibody response of MVC-COV1901 booster was comparable to the response by NVX-CoV2373 booster according to COV-BOOST study [12]. Although all vaccines induced substantial neutralizing antibody titers against alpha and delta variants, significantly lower neutralization against omicron variant was observed in all groups of heterologous vaccine boosting, in line with findings of previous studies [23,24]. The largest reduction was observed in MVC-COV1901. While mRNA booster vaccination could regain the neutralization ability against the wild type and major VOCs, post-booster serum by the protein-based vaccine failed to neutralize the omicron variant. Evidence regarding deficient neutralization against immune-evasion VOCs caused by protein-based vaccine booster remains limited [25]. Why protein-based vaccines failed to stimulate humoral immunity as efficiently as mRNA vaccines is unknown. A development failure in another mRNA vaccine–CVnCoV (CureVac AG) –had been implicated due to the insufficient vaccine dosage design [26]. Therefore, insufficient vaccine antigen may be one of the reasons for the scarcity in neutralization of omicron variant from protein-based vaccine booster.

Accumulated evidence supports that booster vaccination would increase the neutralization capacity against SARS-CoV-2. A previous study showed that in individuals receiving mRNA vaccine one year after natural infection, the vaccination strengthened all components of the B cell response and provoked the serum neutralizing activity against VOCs as well as, or even higher than the wild type strain [27]. Another study also showed similar findings: compared to sera obtained from those two-dose vaccines, sera from individuals after mRNA vaccine booster had a better and more robust correlation of wild-type virus neutralization with delta and omicron, indicating a better cross-neutralization ability [28]. Our study also found that the neutralization titer against alpha and delta were higher than the wild type regardless of the vaccine type, which re-assured the increased cross-neutralization ability after booster vaccination and thereby emphasized the importance of the booster vaccination after emergence of the omicron.

In this study, neutralizing antibody against omicron was under detection level in 60% of participants who received MVC-COV1901 as a booster by LVMNA. However, all participants boosted with MCV-COV1901 had detectable neutralizing antibody against omicron by PNA. Although there is a correlation between LVMNA and PNA, PNA itself could over-express or under-express binding receptors to influence neutralization. On the other hand, the shift of cell entry in the omicron variant may explain this discordance. Omicron has been proved to shift the main cell entry route from the TMPRSS2 dependent pathway to the endocytic pathway, which is significantly different from other VOCs [29□31]. Therefore, the neutralization titer obtained from LVMNA may better reflect omicron’s overall immune evasion performance, including antibody relevant and antibody independent immune evasion.

Although humoral immunity after a booster varied, T cell response against different VOCs remained relatively stable. A study assessing the cross-reactivity of T cell response to the omicron variant showed that T cell reactivity to omicron is preserved in most prior infected and vaccinated individuals [32]. Apart from the primary series, the T cell response after a booster mainly arises from memory T cell proliferation. Our study suggested that all the vaccines used in this study could generate efficient and equal T cell response in HCW primed by the ChAdOx1 nCov-19 in the heterologous booster design.

The study identified no significant difference in neutralizing antibody response between BNT162b2 and half-dose mRNA1273 booster. The effectiveness of the two mRNA vaccines against symptomatic disease by omicron variant in England was also similar [33]. The study revealed that antibody response of half-dose mRNA1273 was in between BNT162b2 and mRNA1273. With less local and systemic adverse events and comparable immunogenicity, the study supports the recommendation to use a half-dose rather than full-dose mRNA1273 as a booster. An earlier study has shown that mRNA vaccines is superior to adenovirus vector vaccines in inducing neutralizing antibody against VOC after booster vaccination [34]. Our study shows that mRNA vaccines are better than a protein vaccine (MVC-COV1901) in neutralizing antibody response to VOCs.

There are limitations of the trial. First, this is not a “mix-and-match” trial. We enrolled only HCWs primed with 2 doses of ChAdOx1 nCov-19 vaccines such that the design cannot evaluate the priming effect of different COVID-19 vaccine. Second, our participants are HCWs, and they are generally young without comorbidities. The immunogenicity in old age, the most vulnerable group, is not available from this study. However, previous studies showed similar booster effect on humoral and cellular responses between younger and older adults [12]. Third, no sophisticated T cell immunity following heterologous booster immunization was performed. Despite that, in a previous immunological study, T cell epitope repertoire was preserved for more than 80% for omicron [35].

The study indicated that LVMNA is a better method to evaluate neutralization activity of immune sera against extremely potent immune evasion omicron variant. Third dose booster not only increases neutralizing antibody titer but also enhance antibody capacity against SARS-CoV-2 variants. Heterologous booster vaccination with mRNA vaccines is recommended for those who have received 2 doses of ChAdOx1 nCov-19.

## Supporting information

Inclusion/Exclusion Criteria

## Data Availability

All data produced in the present study are available upon reasonable request to the authors. All data produced in the present work are contained in the manuscript

## Abbreviations

HCW: health care workers
PNA: pseudovirus neutralizing assay
LVMNA: live virus microneutralization assay
COVID-19: Coronavirus Disease 2019
CGMH: Chang Gung Memorial Hospital
FDA: Food and Drug Administration
VOCs: variants of concern

## Supporting information

**S1 File. CONSORT Checklist**.

**S1 Text. Supporting information_Inclusion/Exclusion Criteria**.

**S2 Text. Supporting information_Methods**.

**S1 Data. Supporting information tables A. Table A**. Severity of adverse events after BNT162b2, half-dose mRNA-1273, mRNA-1273, and MVC-COV1901.

**S2 Data. Supporting information figures A**□**D. Fig A. Local and systemic solicited adverse events of booster vaccines, according to booster regimen**. Shown here are local reactogenicity (Panel A) and systemic reactogenicity (Panel B) that were reported within 1 week after a booster vaccine. The duration of local solicited adverse event was also depicted by weekly interval (Panel C). The severity of adverse event was graded from zero (no symptoms, shown as the lightest color) to four (severely affect daily activity, shown as the darkest color). Grey: BNT162b2; red: half-dose mRNA-1273; yellow: mRNA-1273; blue: MVC-COV1901. **Fig B. Correlation of neutralizing antibodies assessed by correlating binding antibody (Abbott AdviseDx SARS-CoV-2 IgG II assay) to neutralization antibody against D614G and omicron variant (pseudovirus neutralization assay)**. For D614G (left), anti-spike IgG level is highly correlated with PNA (r = 0.87; *P* < 0·0001) but the correlation of both assays is less in omicron (r = 0.59; *P* < 0.0001) (right). The test was performed by Pearson correlation coefficients; *P* < 0.05 was considered statistically significant. **Fig C. Correlation of neutralizing antibodies assessed by live virus microneutralization assay (LVMNA) and pseudovirus neutralization assay (PNA) against Omicron variant**. There is moderate correlation between PNA and LVMNA against omicron variant (r = 0.67, *P* < 0.0001). The test was performed by Pearson correlation coefficient; *P* < 0.05 was considered statistically significant. **Fig D. Correlation of humoral (anti-spike protein binding antibody) and cellular response (interferon-**γ **by ELIspot)**. The humoral response correlated weakly with cellular response in this study (r = 0.17; *P* =0.0016).

## Author Contributions

**Conceptualization:** Chih-Hsien Chuang, Cheng-Hsun Chiu

**Data curation:** Chih-Hsien Chuang, Chung-Guei Huang, Ching-Tai Huang, Yi-Ching Chen, Yu-An Kung, Chih-Jung Chen, Tzu-Chun Chuang, Ching-Chi Liu, Po-Wei Huang, Shu-Li Yang Po-Wen Gu, Shin-Ru Shih

**Formal analysis:** Chih-Hsien Chuang, Chung-Guei Huang, Ching-Tai Huang, Yi-Ching Chen, Yu-An Kung, Chih-Jung Chen, Tzu-Chun Chuang, Ching-Chi Liu

**Funding acquisition:** Shin-Ru Shih, Cheng-Hsun Chiu

**Investigation:** Yu-An Kung, Chih-Jung Chen, Tzu-Chun Chuang, Ching-Chi Liu, Po-Wei Huang, Shu-Li Yang, Po-Wen Gu

**Methodology:** Chih-Hsien Chuang, Chung-Guei Huang, Ching-Tai Huang, Shin-Ru Shih

**Project administration:** Tzu-Chun Chuang, Ching-Chi Liu

**Resources:** Chung-Guei Huang, Ching-Tai Huang, Cheng-Hsun Chiu

**Supervision:** Cheng-Hsun Chiu

**Validation:** Chih-Hsien Chuang, Chung-Guei Huang, Ching-Tai Huang

**Visualization:** Yi-Ching Chen, Tzu-Chun Chuang, Ching-Chi Liu

**Writing – original draft:** Chih-Hsien Chuang, Chung-Guei Huang, Ching-Tai Huang, Yi-Ching Chen, Yu-An Kung, Cheng-Hsun Chiu

**Writing – review & editing:** Shih-Ru Shih, Cheng-Hsun Chiu

## Acknowledgements

We would like to acknowledge the assistance from Clinical Trial Center of Chang Gung Memorial Hospital. Moreover, we would like to acknowledge the volunteers for their willingness to participate in this trial. Additionally, we would like to acknowledge Taiwan Centers for Disease Control and Taiwan Food and Drug Administration, Ministry of Health and Welfare for their scientific input and approval needed to implement this trial. All vaccines were acquired from Taiwan CDC.

## Notes

**Funding:** This work was supported by Medigen Vaccine Biologics Corporation and Chang Gung Memorial Hospital (Dr. Chiu CH, ClinicalTrials.gov number, NCT05132855), under award numbers XMRPG3L0101, CORPG3K0311, CORPG3L0371, CORPG3L0481, and CORPG3K0242. The study was also supported in part by the Research Center for Epidemic Prevention Science, Chang Gung University and Chang Gung Memorial Hospital (award number: MOST 109-2327-B-182-002). The funders had no role in study design, data collection and analysis, decision to publish, or preparation of the manuscript.

### Competing Interest Statement

The authors have declared no competing interest.

### Clinical Trial

ClinicalTrials.gov NCT05132855

### Funding Statement

The trial was sponsored and primarily funded by the Medigen Vaccine Biologics Corporation and Chang Gung Memorial Hospital (both to CHC), under award numbers XMRPG3L0101, CORPG3K0311, CORPG3L0371, CORPG3L0481, and CORPG3K0242 and with support from the Research Center for Epidemic Prevention Science, Chang Gung University and Chang Gung Memorial Hospital (to CHC and Shih SR) (award number: MOST 109-2327-B-182-002). The funders had no role in study design, data collection and analysis, decision to publish, or preparation of the manuscript.

### Author Declarations

The study protocol was approved by Chang Gung Medical Foundation Institutional Review Board (202101767A3).

## References

1. Doria-Rose NA, Shen X, Schmidt SD, O’Dell S, McDanal C, Feng W, et al. Booster of mRNA-1273 strengthens SARS-CoV-2 Omicron neutralization. medRxiv. 2021; 2021.12.15.21267805. https://doi.org/10.1101/2021.12.15.21267805 PMID: 34931200

2. Falsey AR, Frenck RW Jr, Walsh EE, Kitchin N, Absalon J, Gurtman A, et al. SARS-CoV-2 neutralization with BNT162b2 vaccine dose 3. N Engl J Med. 2021; 385: 1627□9. https://doi.org/10.1056/NEJMc2113468 PMID: 34525276

3. Lopez Bernal J, Andrews N, Gower C, Gallagher E, Simmons R, Thelwall S, et al. Effectiveness of Covid-19 vaccines against the B.1.617.2 (Delta) variant. N Engl J Med. 2021; 385: 585□94. https://doi.org/10.1056/nejmoa2108891 PMID: 34289274

4. Greaney AJ, Starr TN, Gilchuk P, Zost SJ, Binshtein E, Loes AN, et al. Complete mapping of mutations to the SARS-CoV-2 spike receptor-binding domain that escape antibody recognition. Cell Host Microbe. 2021; 29: 44□57. https://doi.org/10.1016/j.chom.2020.11.007 PMID: 33259788

5. Rössler A, Riepler L, Bante D, von Laer D, Kimpel J. SARS-CoV-2 Omicron variant neutralization in serum from vaccinated and convalescent persons. N Engl J Med. 2022; 386: 698□700. https://doi.org/10.1056/nejmc2119236 PMID: 35021005

6. Dejnirattisai W, Zhou D, Supasa P, Liu C, Mentzer AJ, Ginn HM, et al. Antibody evasion by the P.1 strain of SARS-CoV-2. Cell 2021; 184: 2939–54.e9. https://doi.org/10.1016/j.cell.2021.03.055 PMID: 33852911

7. Lu L, Mok BW, Chen LL, Chan JM, Tsang OT, Lam BH, et al. Neutralization of SARS-CoV-2 Omicron variant by sera from BNT162b2 or Coronavac vaccine recipients. Clin Infect Dis. 2021. https://doi.org/10.1093/cid/ciab1041 PMID: 34915551.

8. Liu X, Shaw RH, Stuart ASV, Greenland M, Aley PK, Andrews NJ, et al. Safety and immunogenicity of heterologous versus homologous prime-boost schedules with an adenoviral vectored and mRNA COVID-19 vaccine (Com-COV): a single-blind, randomised, non-inferiority trial. Lancet. 2021; 398: 856□69. https://doi.org/10.1016/s0140-6736(21)01694-9 PMID: 34370971

9. Hillus D, Schwarz T, Tober-Lau P, Vanshylla K, Hastor H, Thibeault C, et al. Safety, reactogenicity, and immunogenicity of homologous and heterologous prime-boost immunisation with ChAdOx1 nCoV-19 and BNT162b2: a prospective cohort study. Lancet Respir Med. 2021; 9: 1255□65. https://doi.org/10.1016/s2213-2600(21)00357-x PMID: 34391547

10. Levin EG, Lustig Y, Cohen C, Fluss R, Indenbaum V, Amit S, et al. Waning immune humoral response to BNT162b2 Covid-19 vaccine over 6 months. N Engl J Med 2021; 385: e84. https://doi.org/10.1056/nejmoa2114583 PMID: 34614326

11. Kwok SL, Cheng SM, Leung JN, Leung K, Lee CK, Peiris JM, et al. Waning antibody levels after COVID-19 vaccination with mRNA Comirnaty and inactivated CoronaVac vaccines in blood donors, Hong Kong, April 2020 to October 2021. Euro Surveill. 2022; 27: 2101197. https://doi.org/10.2807/1560-7917.es.2022.27.2.2101197 PMID: 35027105

12. Munro APS, Janani L, Cornelius V, Aley PK, Babbage G, Baxter D, et al. Safety and immunogenicity of seven COVID-19 vaccines as a third dose (booster) following two doses of ChAdOx1 nCov-19 or BNT162b2 in the UK (COV-BOOST): a blinded, multicentre, randomised, controlled, phase 2 trial. Lancet. 2021; 398: 2258□76. https://doi.org/10.1016/s0140-6736(21)02717-3 PMID: 34863358

13. Atmar RL, Lyke KE, Deming ME, Jackson LA, Branche AR, El Sahly HM, et al. Homologous and heterologous Covid-19 booster vaccinations. N Engl J Med. 2022; 386: 1046□57. https://doi.org/10.1056/nejmoa2116414 PMID: 35081293

14. Food and Drug Administration. Toxicity grading scale for health adults and adolescent volunteers enrolled in preventive vaccine clinical trials. Guidance for industry. September 2007. Available from: https://www.fda.gov/media/73679/download access date 22 May 2022

15. Bryan A, Pepper G, Wener MH, Fink SL, Morishima C, Chaudhary A, et al. Performance characteristics of the abbott architect SARS-CoV-2 IgG assay and seroprevalence in Boise, Idaho. J Clin Microbiol. 2020; 58: e00941–20. https://doi.org/10.1128/jcm.00941-20 PMID: 32381641

16. Riester E, Findeisen P, Hegel JK, Kabesch M, Ambrosch A, Rank CM, et al. Performance evaluation of the Roche Elecsys Anti-SARS-CoV-2 S immunoassay. J Virol Methods. 2021; 297: 114271. https://doi.org/10.1016/j.jviromet.2021.114271 PMID: 34461153

17. Chen CY, Liu KT, Shih SR, Ye JJ, Chen YT, Pan HC, et al. Neutralization assessments reveal high cardiothoracic ratio and old age as independent predictors of low neutralizing antibody titers in hemodialysis patients receiving a single dose of COVID-19 Vaccine. J Pers Med. 2022; 12: 68. https://doi.org/10.3390/jpm12010068 PMID: 35055386

18. Le Bert N, Tan AT, Kunasegaran K, Tham CYL, Hafezi M, Chia A, et al. SARS-CoV-2-specific T cell immunity in cases of COVID-19 and SARS, and uninfected controls. Nature. 2020; 584: 457□62. https://doi.org/10.1038/s41586-020-2550-z PMID: 32668444

19. Huang CG, Dutta A, Huang CT, Chang PY, Hsiao MJ, Hsieh YC, et al. Relative COVID-19 viral persistence and antibody kinetics. Pathogens. 2021; 10: 752. https://doi.org/10.3390/pathogens10060752 PMID: 34199240

20. Liu KT, Gong YN, Huang CG, Huang PN, Yu KY, Lee HC, et al. Quantifying neutralizing antibodies in patients with COVID-19 by a two-variable generalized additive model. mSphere. 2022; 7: e0088321. https://doi.org/10.1128/msphere.00883-21 PMID: 35107336

21. Australian Technical Advisory Group on Immunisation (ATAGI) recommendations on the use of a booster dose of COVID-19 vaccine. March 1, 2022. Available from: https://www.health.gov.au/sites/default/files/documents/2022/03/atagi-recommendations-on-the-use-of-a-booster-dose-of-covid-19-vaccine.pdf access date 25 May 2022

22. Sablerolles RSG, Rietdijk WJR, Goorhuis A, Postma DF, Visser LG, Geers D, et al. Immunogenicity and reactogenicity of vaccine boosters after Ad26.COV2.S priming. N Engl J Med. 2022; 386: 951□63. https://doi.org/10.1056/nejmoa2116747 PMID: 35045226

23. Pajon R, Doria-Rose NA, Shen X, Schmidt SD, O’Dell S, McDanal C, et al. SARS-CoV-2 Omicron variant neutralization after mRNA-1273 booster vaccination. N Engl J Med. 2022; 386: 1088□91. https://doi.org/10.1056/nejmc2119912 PMID: 35081298

24. Nemet I, Kliker L, Lustig Y, Zuckerman N, Erster O, Cohen C, et al. Third BNT162b2 vaccination neutralization of SARS-CoV-2 Omicron infection. N Engl J Med. 2022; 386: 492□4. https://doi.org/10.1056/nejmc2119358 PMID: 34965337

25. Ai J, Zhang H, Zhang Y, Lin K, Zhang Y, Wu J, et al. Omicron variant showed lower neutralizing sensitivity than other SARS-CoV-2 variants to immune sera elicited by vaccines after boost. Emerg Microbes Infect. 2022; 11: 337□3. https://doi.org/10.1080/22221751.2021.2022440 PMID: 34935594

26. Dolgin E. CureVac COVID vaccine let-down spotlights mRNA design challenges. Nature. 2021; 594: 483. https://doi.org/10.1038/d41586-021-01661-0 PMID: 34145413

27. Wang Z, Muecksch F, Schaefer-Babajew D, Finkin S, Viant C, Gaebler C, et al. Naturally enhanced neutralizing breadth against SARS-CoV-2 one year after infection. Nature. 2021; 595: 426□31. https://doi.org/10.1038/s41586-021-03696-9 PMID: 34126625

28. Garcia-Beltran WF, St Denis KJ, Hoelzemer A, Lam EC, Nitido AD, Sheehan ML, et al. mRNA-based COVID-19 vaccine boosters induce neutralizing immunity against SARS-CoV-2 Omicron variant. Cell. 2022; 185: 457□66.e4. https://doi.org/10.1016/j.cell.2021.12.033 PMID: 34995482

29. Pia L, Rowland-Jones S. Omicron entry route. Nat Rev Immunol. 2022; 22: 144. https://doi.org/10.1038/s41577-022-00681-9 PMID: 35082449

30. Hui KPY, Ho JCW, Cheung MC, Ng KC, Ching RHH, Lai KL, et al. SARS-CoV-2 Omicron variant replication in human bronchus and lung ex vivo. Nature. 2022; 603: 715□20. https://doi.org/10.1038/s41586-022-04479-6 PMID: 35104836

31. Meng B, Abdullahi A, Ferreira IATM, Goonawardane N, Saito A, Kimura I, et al. Altered TMPRSS2 usage by SARS-CoV-2 Omicron impacts infectivity and fusogenicity. Nature. 2022; 603: 706□14. https://doi.org/10.1038/s41586-022-04474-x PMID: 35104837

32. Naranbhai V, Nathan A, Kaseke C, Berrios C, Khatri A, Choi S, et al. T cell reactivity to the SARS-CoV-2 Omicron variant is preserved in most but not all individuals. Cell. 2022; 185: 1041□51.e6. https://doi.org/10.1101/2022.01.04.21268586 PMID: 35018386

33. Andrews N, Stowe J, Kirsebom F, Toffa S, Rickeard T, Gallagher E, et al. Covid-19 vaccine effectiveness against the Omicron (B.1.1.529) variant. N Engl J Med. 2022: 386: 1532□46. https://doi.org/10.1056/nejmoa2119451 PMID: 35249272

34. van Gils MJ, Lavell A, van der Straten K, Appelman B, Bontjer I, Poniman M, et al. Antibody responses against SARS-CoV-2 variants induced by four different SARS-CoV-2 vaccines in health care workers in the Netherlands: A prospective cohort study. PLoS Med. 2022; 19: e1003991. https://doi.org/10.1371/journal.pmed.1003991 PMCID: 35580156

35. Tarke A, Coelho CH, Zhang Z, Dan JM, Yu ED, Methot N, et al. SARS-CoV-2 vaccination induces immunological T cell memory able to cross-recognize variants from Alpha to Omicron. Cell. 2022; 185: 847□59.e11. https://doi.org/10.1016/j.cell.2022.01.015 PMID: 35139340

