## SupplementaL Table A for "Titers and capacity of neutralizing antibodies against SARS-CoV-2 variants after heterologous booster vaccination in health care workers primed with two doses of ChAdOx1 nCov-19: a single-blinded, randomized clinical trial"

| **S1 Data_Supporting information_tables A**  **Table A. Severity of Adverse Events After BNT162b2, Half-dose mRNA-1273, mRNA-1273, and MVC-COV1901.** | | | | | | |
| --- | --- | --- | --- | --- | --- | --- |
| **Side effects in 7 consecutive days after booster vaccines** | | **BNT162b2** | **Half-dose**  **mRNA-1273** | **mRNA-1273** | **MVC-COV1901** | ***P* value** |
| Pain | No at all | 3 (3.6%) | 3 (3.6%) | 0 (0%) | 20 (23%) | *P* < 0.0001 |
|  | Dose not interference with activity | 74 (89.2%) | 64 (75.3%) | 61 (71.8%) | 65 (76.5%) |  |
|  | Repeated use of non-narcotic pain reliever >24 hours or interferes with activity | 6 (7.2%) | 18 (21.2%) | 22 (25.9%) | 0 (0%) |  |
|  | Any use of narcotic pain reliever or prevents daily activity | 0 (0%) | 0 (0%) | 2 (2.4%) | 0 (0%) |  |
| Erythema/ Redness at injection site | Not at all | 74 (89.2%) | 70 (82.4%) | 68 (80%) | 83 (97.6%) | *P* = 0.0050 |
|  | 2.5-5 cm | 8 (9.6%) | 8 (9.4%) | 12 (14.1%) | 2 (2.4%) |  |
|  | 5-10 cm | 1 (1.2%) | 7 (8.2%) | 5 (5.9%) | 0 (0%) |  |
|  | >10 cm | 0 (0%) | 0 (0%) | 0 (0%) | 0 (0%) |  |
| Swelling | Not at all | 15 (18.1%) | 11 (12.9%) | 4 (4.7%) | 36 (42.4%) | *P* < 0.0001 |
|  | Dose not interferes with activity | 58 (69.9%) | 53 (62.4%) | 55 (64.7%) | 46 (54.1%) |  |
|  | Interferes with activity | 10 (12.0%) | 20 (23.5%) | 26 (30.6%) | 3 (3.5%) |  |
|  | Prevents daily activity | 0 (0%) | 0 (0%) | 0 (0%) | 0 (0%) |  |
| Fever | Not at all | 74 (89.2%) | 72 (84.7%) | 60 (70.6%) | 81 (95.3%) | *P* < 0.0001 |
|  | 38.0-38.4 | 2 (2.4%) | 11 (12.9%) | 14 (16.5%) | 2 (2.4%) |  |
|  | 38.5-38.9 | 5 (6.0%) | 2 (2.4%) | 6 (7.1%) | 2 (2.4%) |  |
|  | 39.0-40.0 | 2 (2.4%) | 0 (0%) | 5 (5.9%) | 0 (0%) |  |
| Fatigue | Not at all | 21 (25.3%) | 26 (30.6%) | 4 (4.7%) | 40 (47.1%) | *P* < 0.0001 |
|  | Little, no interference with activity | 40 (48.2%) | 30 (35.3%) | 49 (57.6%) | 40 (47.1%) |  |
|  | Some interference with activity | 20 (24.1%) | 29 (34.1%) | 28 (32.9%) | 4 (4.7%) |  |
|  | Significant; prevents daily activity | 2 (2.4%) | 0 (0%) | 4 (4.7%) | 1 (1.2%) |  |
| Myalgia | Not at all | 26 (31.3%) | 22 (25.9%) | 10 (11.8%) | 54 (63.5%) | *P* < 0.0001 |
|  | Little, no interference with activity | 51 (61.4%) | 49 (57.7%) | 54 (63.5%) | 31 (36.5%) |  |
|  | Some interference with activity | 6 (7.2%) | 14 (16.5%) | 20 (23.5%) | 0 (0%) |  |
|  | Significant; prevents daily activity | 0 (0%) | 0 (0%) | 1 (1.2%) | 0 (0%) |  |
| Headache | Not at all | 47 (56.6%) | 41 (48.2%) | 28 (32.9%) | 53 (62.4%) | *P* = 0.0003 |
|  | Little, no interference with activity | 32 (38.6%) | 30 (35.3%) | 41 (48.2%) | 31 (36.5%) |  |
|  | Repeated use of non-narcotic pain reliever >24 hours or some interference with activity | 4 (4.8%) | 13 (15.3%) | 16 (18.8%) | 1 (1.2%) |  |
|  | Significant; any use of narcotic pain reliever or prevents daily activity | 0 (0%) | 1 (1.2%) | 0 (0%) | 0 (0%) |  |
| Nausea/ Vomiting | Not at all | 65 (78.3%) | 69 (81.2%) | 61 (71.8%) | 73 (85.9%) | *P* = 0.44 |
|  | Little, no interference with activity or 1-2 episodes/24 hours | 15 (18.1%) | 12 (14.1%) | 19 (22.4%) | 10 (11.8%) |  |
|  | Some interference with activity >2 episodes/24 hours | 3 (3.6%) | 4 (4.7%) | 5 (5.9%) | 2 (2.4%) |  |
|  | Prevents daily activity, requires outpatient IV hydration | 0 (0%) | 0 (0%) | 0 (0%) | 0 (0%) |  |
| Diarrhea | Not at all | 69 (83.1%) | 66 (77.6%) | 77 (90.6%) | 77 (90.6%) | *P* = 0.18 |
|  | 2-3 loose stools or <400 gms/24 hours | 13 (15.7%) | 18 (21.2%) | 7 (8.2%) | 8 (9.4%) |  |
|  | 4-5 loose stools or <400-800 gms/24 hours | 1 (1.2%) | 1 (1.2%) | 1 (1.2%) | 0 (0%) |  |
|  | 6 or more watery stools or >800 gms/24 hours or requires outpatient IV hydration | 0 (0%) | 0 (0%) | 0 (0%) | 0 (0%) |  |
