## Supplementary material for "Titers and capacity of neutralizing antibodies against SARS-CoV-2 variants after heterologous booster vaccination in health care workers primed with two doses of ChAdOx1 nCov-19: a single-blinded, randomized clinical trial": Inclusion/Exclusion Criteria

**S1 Text_Supporting information_ Inclusion/Exclusion Criteria**

**Inclusion criteria**

1. Participants willing to give written informed consent in the trial will be qualified for participation by infectious disease physicians at the 1^st^ visit.

2. Age between 20 to 65 years old.

3. Already received 2 doses of the ChAdOx1 for more than 90 days. Evidence of this will be gathered from medical history and/or medical records including the COVID-19 vaccine registration yellow card.

**Exclusion criteria**

The participant may not enter the trial if ANY of the following apply:

1. Fever or evidence of upper respiratory tract infections
2. Confirmed COVID-19 cases (PCR-confirmed infection or detectable anti-nucleocapsid protein IgG)
3. History of anaphylaxis, severe allergic disease or reactions likely to be exacerbated by any component of study vaccines (e.g. hypersensitivity to the active substance or any of the listed ingredients of any study vaccine).
4. Malignancy requiring receipt of immunosuppressive chemotherapy or radiotherapy for treatment of solid organ cancer/hematological malignancy within the 6 months prior to enrollment.
5. Bleeding disorder (e.g. factor deficiency, coagulopathy or platelet disorder), or prior history of significant bleeding or bruising following intramuscular injections or venipuncture.
6. Has received vaccines other than COVID-19 vaccine within one month
7. Pregnancy or willingness/intention to become pregnant within 3 months post booster vaccine
8. Aged < 20 years or unable to sign informed consent
9. Any other significant disease, disorder or finding which may significantly increase the risk to the volunteer because of participation in the study, affect the ability of the volunteer to participate in the study or impair interpretation of the study data or insufficient level of language to undertake all study requirements in opinion of the Investigators.
10. Blood or liver dysfunction determined by blood tests at the 1^st^ visit, including CBC/DC and AST、ALT, determined by infectious disease physician.
11. Female subjects aged 20-40 years old with positive hCG pregnancy test (human chorionic gonadotropin urine test)
12. If the anti-N antibody test is positive from the blood test at the 1^st^ visit, the case will be excluded from the analysis of immunogenicity effectiveness in the follow-up.
