## Supplemental Figures for "Titers and capacity of neutralizing antibodies against SARS-CoV-2 variants after heterologous booster vaccination in health care workers primed with two doses of ChAdOx1 nCov-19: a single-blinded, randomized clinical trial"

**S2 Data_Supporting information_figures A-D**

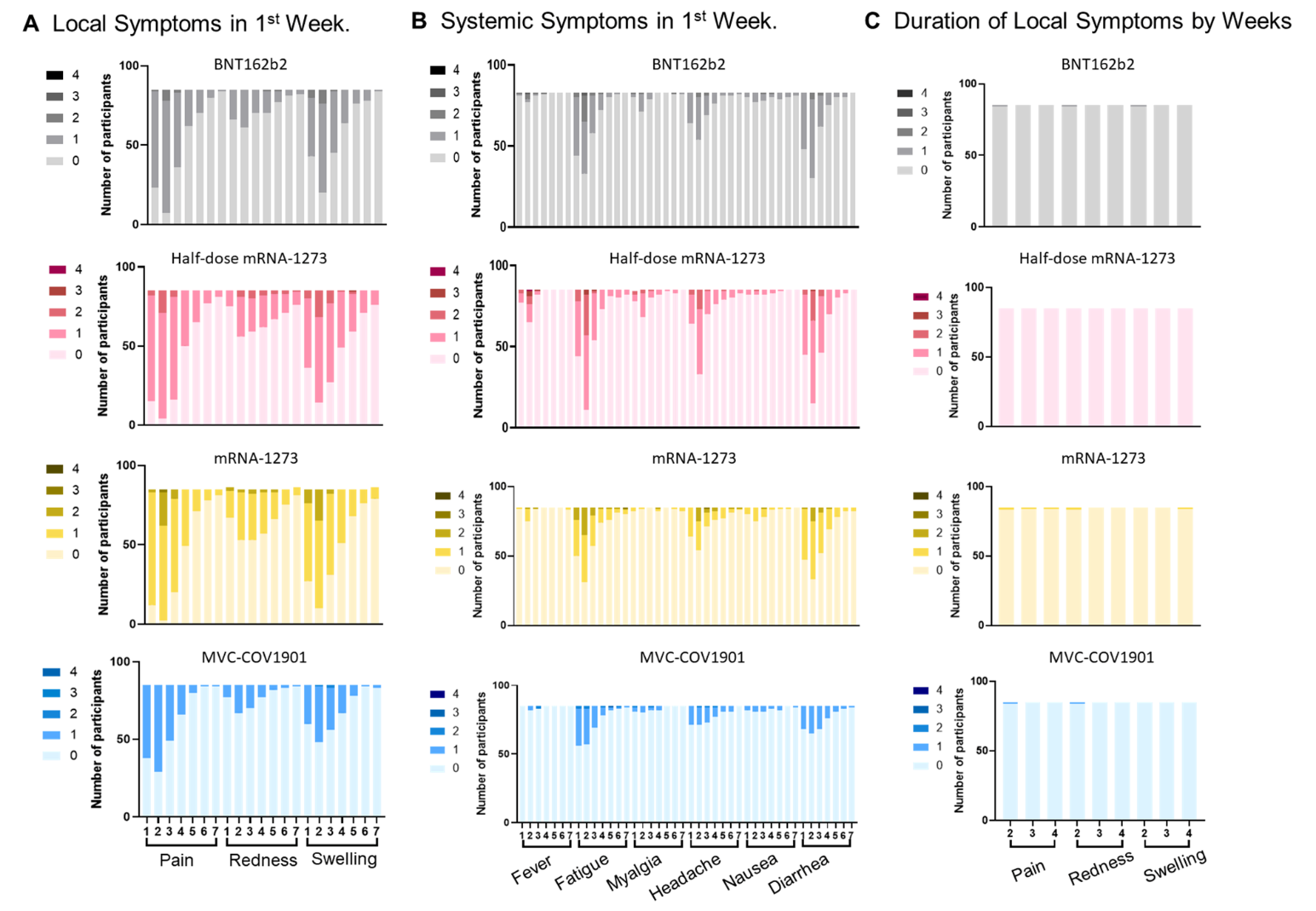

**Fig A. Local and systemic solicited adverse events of booster vaccines, according to booster regimen.** Shown are local reactogenicity (Panel A) and systemic reactogenicity (Panel B) that were reported within 1 week after a booster vaccine. The duration of local solicited adverse event which was depicted by week interval was shown in panel C. The severity of adverse event was graded from zero (no symptoms, shown as the lightest color) to four (severely affect daily activity, shown as the darkest color). Grey: BNT162b2; red: half-dose mRNA-1273; yellow: mRNA-1273; blue: MVC-COV1901.

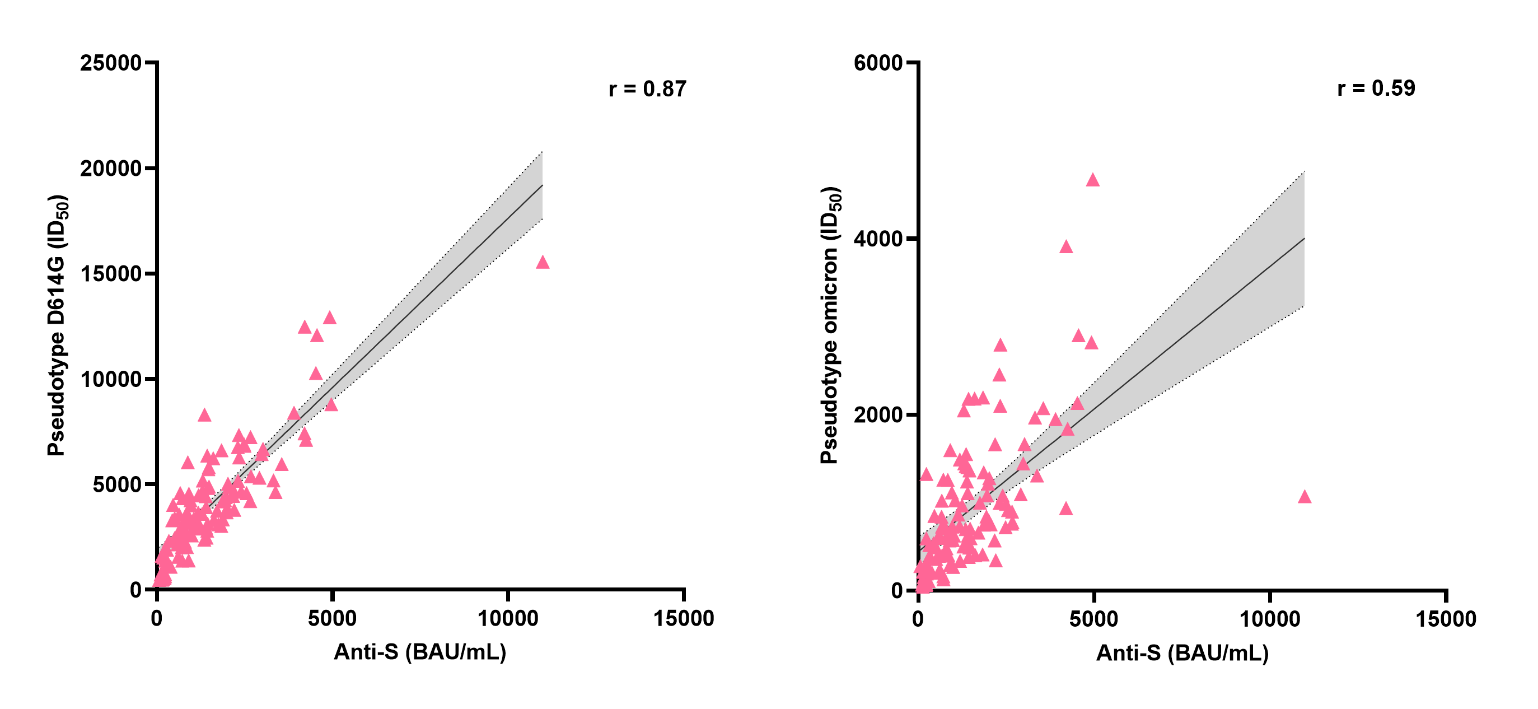

**Fig B. Correlation of Binding Antibody (Abbott AdviseDx SARS-CoV-2 IgG II assay) and Pseudovirus Neutralization Assay (PNA) against D614G and Omicron variant.** For D614G (left), anti-spike IgG level is highly correlated with PNA (*r* = 0.87; P < 0.0001) but the correlation of both assays is less in omicron (*r* = 0.59; P < 0.0001) (right). The test was performed by Pearson correlation coefficient; *P* < 0.05 was considered statistically significant.

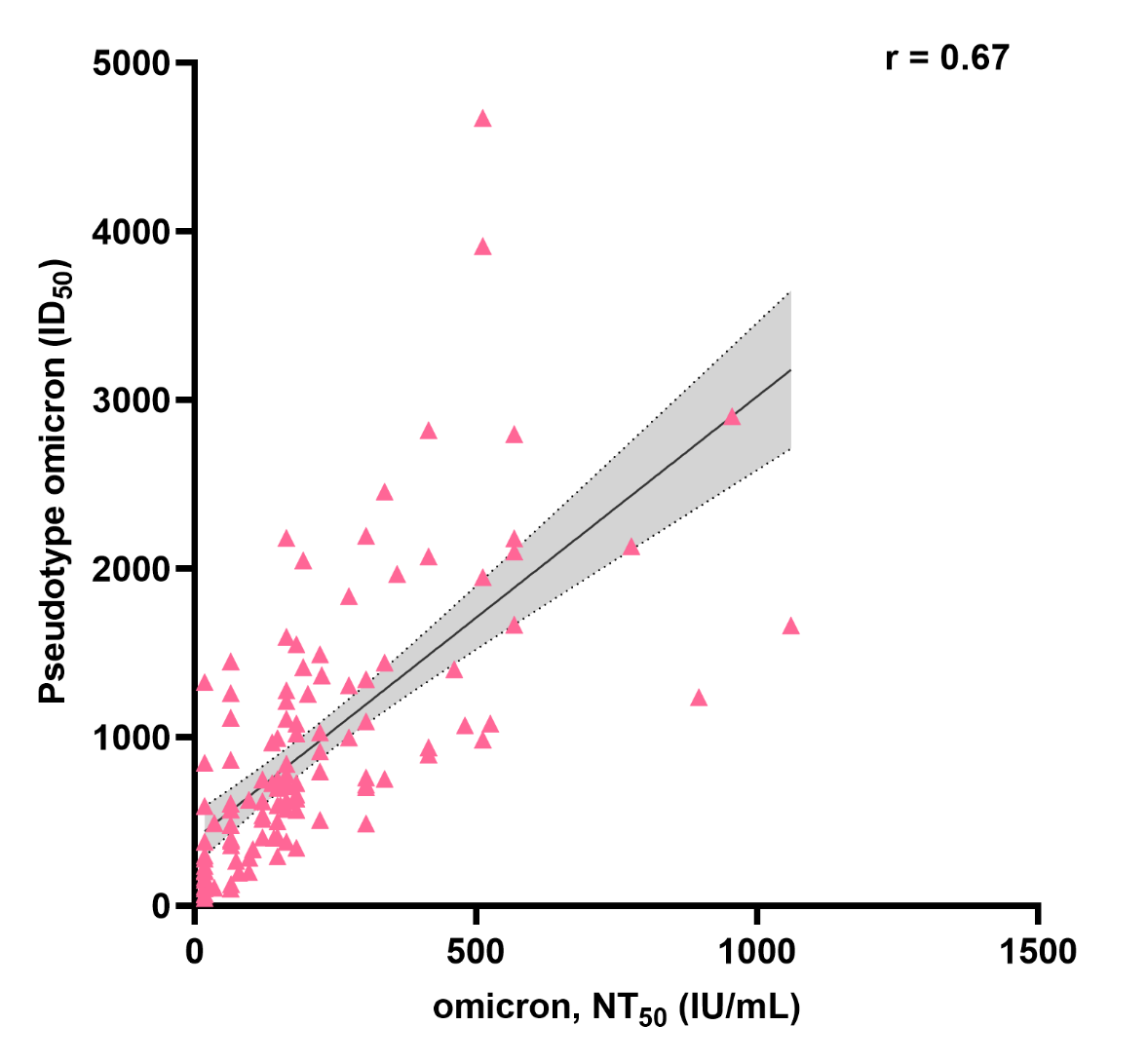

**Fig C.** **Correlation of neutralizing antibodies accessing by Live Virus Microneutralization Assay (LVMNA) and Pseudovirus Neutralization Assay (PNA) against Omicron variant.**  There is moderate correlation between PNA and LVMNA  against omicron variant (r= 0.67, P < 0.0001). The test was performed by Pearson correlation coefficient; P < 0.05 was considered statistically significant.

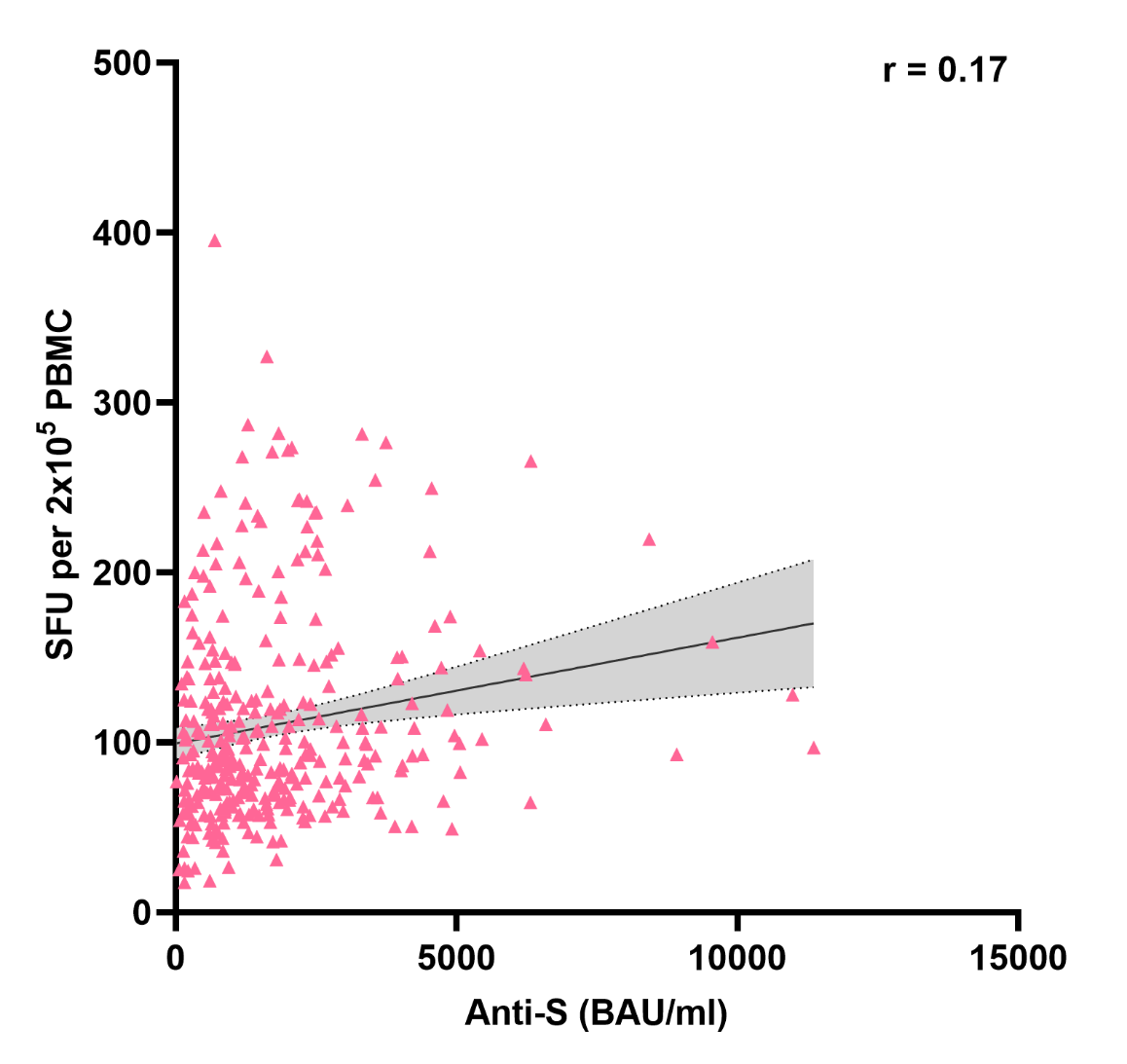

**Fig D.** C**orrelation of humoral (anti-spike protein binding antibody) and cellular responses (ELIspot).** The humoral response correlated weakly with cellular responses in our study (r = 0.17; P = 0.0016).
