## Supplemental Methods for "Titers and capacity of neutralizing antibodies against SARS-CoV-2 variants after heterologous booster vaccination in health care workers primed with two doses of ChAdOx1 nCov-19: a single-blinded, randomized clinical trial"

**Supporting information_Methods**

**Serologic Assays**

Each serum sample was analyzed by the MeDiPro SARS-CoV-2 antibody ELISA (cat No. IR-0061E), Abbott AdviseDx SARS-CoV-2 IgG II assay (cat No. 6S61.22), and Roche Elecsys Anti-SARS-COV-2 (Cat. 09203079190) according to the manufacturers’ instructions [1‒3]. The MeDiPro SARS-CoV-2 antibody ELISA detected antibodies against S1 and RBD, and values <12 IU/mL were considered negative. The Abbott AdviseDx SARS-CoV-2 IgG II assay is a chemiluminescent microparticle immunoassay (CMIA) for the detection of IgG antibodies to the RBD of S protein; the cut-off value was 50.0 AU/mL. The Abbott AU/mL was transferred to WHO unit (binding antibody unit per mL (BAU/mL)) followed the equation of 1 AU/mL = 0.142 BAU/mL. The Roche Elecsys Anti-SARS-COV-2 assay used is an electro-chemiluminescence immunoassay (ECLIA) for the detection of IgG antibodies to the Nucleocapsid (N) protein; the cut-off value was 1.0 COI.

**Quantifying Neutralization Antibodies (nAbs) by a Two-Variable Generalized Additive Model (ELISA NT)**

The MeDiPro SARS-CoV-2 Antibody ELISA kit was designed to detect SARS-CoV-2 nAbs in the serum, based on the binding affinity of S1 and RBD to antibodies. The RBD is the major binding site of nAbs. S1 covers the RBD and several other regions, which are also imperative for nAb binding. The assay combines each of the S1 and RBD ELISA unit (EU) values and applies spline-based generalized additive model (GAM) regression analysis (using S1 and RBD as two predictors) to predict NT_50_ by combining multiple smooth functions. MeDiPro is a kit for quantifying nAbs using technology transferred from the Research Center for Emerging Viral Infections, Chang Gung University, and has been approved by the Taiwan Food and Drug Administration (No. 1106803303); the data for S1 and RBD fusion proteins can accurately predict the SARS-CoV-2 50% neutralization titer (NT50) under a two-variable generalized additive model and WHO international unit conversion (Supplementary Materials). According to the manufacturer, this test has 92.2% (95% CI, 84.0–96.4%) sensitivity and 93% (95% CI, 81.4–97.6%) specificity.

**Live Virus Microneutralization Assay**

The following SARS-COV-2 viral strains isolated in Chang Gung Memorial Hospital were used for BSL3 neutralizing antibody testing: wild type strain (reference sequence ID_ MT192759 ), alpha (B.1.1.7, reference sequence ID_ MZ277391 ), delta (B.1.617.2, reference sequence ID_ON005386 ), and omicron (B.1.1.529, reference sequence ID_ON005319 ). We followed the standard protocol of a plaque reduction neutralization test [4]. Vero E6 cells (2 × 10^4^ cells per well) were seeded in a 96-well plate and incubated at 37°C for 24 h. The medium was replaced with 100 μL of fresh DMEM containing 2% FBS. The live virus microneutralization assay was performed in a BSL-3 laboratory using the quantified SARS-CoV-2 viral strain. All serum samples were heat-inactivated at 56°C for 30 min and then 2-fold serially diluted in DMEM (Gibco) without FBS. From a starting dilution of 1:8 for each sample, ten 2-fold dilutions were performed for a final dilution of 1:8192. Each serum sample was incubated with 100 50% tissue culture infectious doses (100 TCID50) of SARS-CoV-2 at 37°C for 1 h prior to infection with Vero E6 cells. Add 100 μL of the virus-serum mixtures at each dilution to a 96-well plate containing the confluent Vero E6 monolayer. After infected cells were incubated at 37°C for 5 days, they were fixed with 10% formaldehyde and stained with crystal violet. The neutralization titer was calculated as the logarithm of the 50% end point using the Reed–Muench method based on the presence or absence of cytopathic effects. Each serum sample was tested in four replicates. To convert the NT_50_ to the World Health Organization (WHO) international standard unit (IU), WHO IS sera (NIBSC code 20/136) and WHO Reference Panel for anti-SARS-CoV-2 antibody (NIBSC code 20/268) were obtained from National Institute for Biological Standards and Control (NIBSC) [5]. Each standard serum has its own IU value. The NT_50_ values for WHO IS and Reference Panel sera were determined by a live virus microneutralization assay. Based on the NT_50_ and IU values for WHO IS and Reference Panel sera, the calibration curve (standard curve) was established by a simple linear regression model used for the conversion of NT_50_ values to the international standard units (IU/mL).

**SARS-CoV-2 Pseudovirus Neutralization Assay [6]**

SARS-CoV-2 pseudovirus expressing the wild type (D614G) or omicron spike protein were prepared and titrated by the National RNAi Core Facility, Academia Sinica, Taiwan. 293T-ACE2 cells were seed at 4 × 10^4^ cells per well in a 96-well plate and incubated at 37°C for 24 h. The serum samples from the vaccinated individuals began to neutralize at an 8-fold dilution, and then were serially diluted 2-fold in DMEM without FBS. The diluted serum samples were incubated with SARS-CoV-2 pseudovirus (6 × 10^3^ relative infection units) for 1 h at 37°C. After incubation, 293T-ACE2 cells were infected with diluted serum and SARS-CoV-2 pseudovirus in duplicate for 24 h at 37°C, and then the medium was removed and replaced with DMEM with 10% FBS for an additional 24 h incubation. Finally, the medium was removed by gentle aspiration and 100 µl of Bright-Glo luciferase reagent (Promega, Madison, WI, USA) was added to all wells. The luminescence signal was measured using a Synergy 2 microplate reader. SARS-CoV-2 neutralizing antibody titer was expressed as the reciprocal of serum samples at which RLU were reduced by 50% dilution (ID50) compared to the mean virus control wells. ID50 values were calculated in GraphPad Prism 8. The limit of detection of this assay is an ID50 of 8.

**T Cell ELISpot**

SARS-CoV-2-specific T cell responses were detected using the Human IFN-gamma ELISpot Kit (EL285, R&D) according to the manufacturer’s instructions [7]. In brief, peripheral blood mononuclear cells (PBMCs) were isolated from whole blood samples, diluted with PBS, is gently layered over an equal volume of Ficoll-Paque PLUS (17144003, Cytiva) in a 50-ml tube then centrifuged for 20 minutes at 2000 rpm. In the 96-well PVDF-backed microplate coated with a monoclonal antibody specific for human IFN-γ, 100 µl of 2.5 x 10^5^ PBMCs were stimulated with 1 µg of Recombinant SARS-CoV-2 Spike His Protein (10549-CV-01M, R&D) and positive (phytohaemagglutinin, PHA, 10576015, Thermo Fisher) and negative controls. Cells were incubated in a humidified 37℃ CO_2_ for 16-22hr and interferon-γ secreting T cells were detected. Spot-forming units (SFUs) were detected using an automated plate reader (CTL ImmunoSPOT Analyzer).

**References**

1. Bryan A, Pepper G, Wener MH, Fink SL, Morishima C, Chaudhary A, Jerome KR, et al. Performance characteristics of the Abbott architect SARS-CoV-2 IgG assay and seroprevalence in Boise, Idaho. J Clin Microbiol. 2020; 58: e00941-20. <https://doi.org/10.1128/jcm.00941-20> PMID: [32381641](https://pubmed.ncbi.nlm.nih.gov/32381641)
2. Riester E, Findeisen P, Hegel JK, et al. Performance evaluation of the Roche Elecsys Anti-SARS-CoV-2 S immunoassay. J Virol Methods. 2021; 297: 114271. <https://doi.org/10.1016/j.jviromet.2021.114271> PMID: [34461153](https://pubmed.ncbi.nlm.nih.gov/34461153)
3. Chen CY, Liu KT, Shih SR, Ye JJ, Chen YT, Pan HC,et al. Neutralization assessments reveal high cardiothoracic ratio and old age as independent predictors of low neutralizing antibody titers in hemodialysis patients receiving a single dose of COVID-19 Vaccine. J Pers Med. 2022; 12: 68. <https://doi.org/10.3390/jpm12010068> PMID: [35055386](https://pubmed.ncbi.nlm.nih.gov/35055386)
4. Huang CG, Dutta A, Huang CT, Chang PY, Hsiao MJ, Hsieh YC, et al. Relative COVID-19 viral persistence and antibody kinetics. Pathogens. 2021; 10: 752. <https://doi.org/10.3390/pathogens10060752> PMID: [34199240](https://pubmed.ncbi.nlm.nih.gov/34199240)
5. https://cdn.who.int/media/docs/default-source/biologicals/ecbs/bs-2020-2403-sars-cov-2-ab-ik-17-nov-2020_4ef4fdae-e1ce-4ba7-b21a-d725c68b152b.pdf
6. Liu KT, Gong YN, Huang CG, Huang PN, Yu KY, Lee HC, et al. Quantifying neutralizing antibodies in patients with COVID-19 by a two-variable generalized additive model. mSphere. 2022; 7: e0088321. <https://doi.org/10.1128/msphere.00883-21> PMID: [35107336](https://pubmed.ncbi.nlm.nih.gov/35107336)
7. Le Bert N, Tan AT, Kunasegaran K, Tham CYL, Hafezi M, Chia A, et al. SARS-CoV-2-specific T cell immunity in cases of COVID-19 and SARS, and uninfected controls. Nature. 2020; 584: 457‒62. <https://doi.org/10.1038/s41586-020-2550-z> PMID: [32668444](https://pubmed.ncbi.nlm.nih.gov/32668444/)
